# Enhanced health facility surveys to support malaria control and elimination across different transmission settings in The Philippines

**DOI:** 10.1101/2020.07.07.20146225

**Authors:** Ralph A. Reyes, Kimberly M. Fornace, Maria Lourdes M. Macalinao, Beaulah L. Boncayao, Ellaine S. De La Fuente, Hennessey M. Sabanal, Alison Paolo N. Bareng, Inez Andrea P. Medado, Edelwisa S. Mercado, Jennifer S. Luchavez, Julius Clemence R. Hafalla, Chris J. Drakeley, Fe Esperanza J. Espino

**Affiliations:** Department of Parasitology, Research Institute for Tropical Medicine, 9002 Research Drive, Filinvest Corporate City, Alabang, Muntinlupa City, 1781, Metro Manila, Philippines; Faculty of Infectious and Tropical Diseases, London School of Hygiene and Tropical Medicine, Keppel Street, London WC1E 7HT, United Kingdom; Molecular Biology Laboratory, Research Institute for Tropical Medicine, 9002 Research Drive, Filinvest Corporate City, Alabang, Muntinlupa City, 1781, Metro Manila, Philippines

## Abstract

Following substantial progress in malaria control in the Philippines, new surveillance approaches are needed to identify and target residual malaria transmission. This study evaluated an enhanced surveillance approach using rolling cross-sectional surveys of all health facility attendees augmented with molecular diagnostics and geolocation. Facility surveys were carried out in 3 sites representing different transmission intensities: Morong, Bataan (pre-elimination), Abra de Ilog, Occidental Mindoro (stable-medium risk) and Rizal, Palawan (high risk, control). Only 1 RDT positive infection and no PCR confirmed infections were found in Bataan and Occidental Mindoro suggesting the absence of transmission. In Rizal, inclusion of all health facility attendees, regardless of symptoms, and use of molecular diagnostics identified an additional 313 infected individuals in addition to 300 cases identified by routine screening of febrile patients with RDT or microscopy. Of these, the majority (313/613) were subpatent infections and only detected using molecular methods. Simultaneous collection of GPS coordinates on tablet-based applications allowed real-time mapping of malaria infections. Risk factor analysis showed higher risks in children and indigenous groups, with bednet use having a protective effect. Subpatent infections were more common in men and older age groups. Overall, malaria risks were not associated with patient status and some of non-patient clinic attendees reported febrile illnesses (1.9%, 26/1369) despite not seeking treatment highlighting the widespread distribution of infection in communities. Together, these data illustrate the utility of health-facility based surveys to augment surveillance data to increase the probability of detecting infections in the wider community.

## Background

The Philippines declared its vision of eliminating malaria by 2030 with a goal of reducing malaria incidence in the country by 90% relative to a 2016 baseline of 6,604 reported cases. Through its strategy of sub-national elimination, enhanced case detection and treatment and vector control, aims to increase the number of malaria free provinces from 32 to 74 by 2022 out of the 81 provinces ^1, 2^. However, malaria continues to be a public health burden with highly variable transmission across the country. In 2018, 4,902 indigenous cases and 1 death were reported with approximately 95% of these on Palawan island (API ≥ 1 per 1,000 at-risk population). Within Palawan, transmission is geographically heterogeneous, with malaria free municipalities in the north and southern municipalities endemic for all five human *Plasmodium* species.

Recent World Health Organization (WHO) guidelines on malaria surveillance define surveillance as a core intervention required in settings of any level of transmission to meet elimination goals. The guidelines also highlighted the need for increasingly spatially and temporally resolute data on malaria infection as transmission declines ^3^. While population-based community surveys remain the gold standard for measuring prevalence and assessing spatial patterns of infection, these sampling approaches are highly resource intensive and may require prohibitively large sample sizes in low transmission settings. Alternatively, surveys of easy access groups, such as health facility attendees or school children, can be used to provide rapid estimates of malaria prevalence within the community (e.g. ^4-8^). These surveys may not fully capture the distribution of infection in the entire population but are operationally feasible and cost effective to implement. As malaria transmission decreases, spatial heterogeneity becomes more pronounced, with substantial variations observed in the geographic distribution of infections ^9^. However, by incorporating methods of geolocating participant households using tablet-based applications, fine-scale maps of malaria infection can be created in near real-time, allowing identification of foci of transmission ^10^ which are relevant for areas like Palawan.

Additionally, conventional diagnostic methods recommended by the WHO have limitations for surveillance as low parasite density resulting to submicroscopic and asymptomatic infections are missed ^11, 12^. With only symptomatic infections being tested, individuals who are not seeking treatment are overlooked and malaria transmission estimates based on clinical cases reporting to health facilities are biased ^13^. Asymptomatic and subpatent infections comprise the majority of malaria infections in low endemic areas despite adequate malaria control measures and contribute to maintaining transmission, undermining elimination efforts ^14^. Most of these infections are not detectable by conventional microscopy or rapid diagnostic tests (RDTs), necessitating the use of molecular techniques ^15, 16^. Detecting these infections can be challenging due to the infrequent reports of clinical cases and low probability of identifying infections.

To assess how health facility-based surveys with molecular diagnostics could be utilized to support malaria elimination efforts, we conducted rolling cross-sectional surveys in the provinces of Palawan, Occidental Mindoro and Bataan, three areas of the Philippines with different levels of reported transmission. The overall aims were to (1) develop methods for health facility-based surveys applying improved diagnostics and geolocation technologies, (2) assess the utility of enhanced surveillance approaches to improve detection of malaria infections and (3) identifying characteristics of individuals with subpatent infections.

## Methods

### Study areas

The areas were selected based on the 2014 Philippines’ National Malaria Program operational definition of malaria endemic provinces (Figure 1). In that year, Palawan, Occidental Mindoro, and Bataan were categorized as stable-high risk or control phase, stable-medium risk of transmission and malaria pre-elimination provinces, respectively, and in the same order, the annual parasitological indices were 0, 0.35 and 5.7 respectively in 2018 ^17^. Demographic information and land areas of the selected areas are shown in Table 1. According to Philippine Statistics Authority Census of 2000, the population in all study sites is comprised of various ethnicities and indigenous groups. In Palawan, almost half of the population belong to different indigenous groups. The Palaw’an indigenous group comprise 38.7% of the total population in Rizal; other ethnic groups include Kagayanen (1.9%), Tausug (1.8%), Cuyunon (1.2%), Maranao (1.1%), Jama Mapun (0.9%) and Agutaynen (0.1%) ^18^. Main occupations include subsistence farmers, swidden agriculture and fisherman. In Abra de Ilog, Occidental Mindoro, 64.2% of the population comprises Tagalog and 30.6% of indigenous groups while majority of the population in Morong, Bataan classified themselves as Tagalog (91.0%) with 0.8% of indigenous population ^19, 20^. Residents in Abra de Ilog and Morong are primarily long-time settlers with small businesses. All provinces are predominantly rural, partly forested with seasonal rainfall generally from May to October. Primary health care services are provided by the rural health unit (RHU) and barangay health stations. In addition to these facilities and to service remote communities, Abra de Ilog and Rizal have malaria testing (using RDTs) and treatment centers based at households of community health workers. With supervision from RHU staff, community volunteers operate the barangay health stations and remote malaria testing and treatment centers.

**Table 1.**
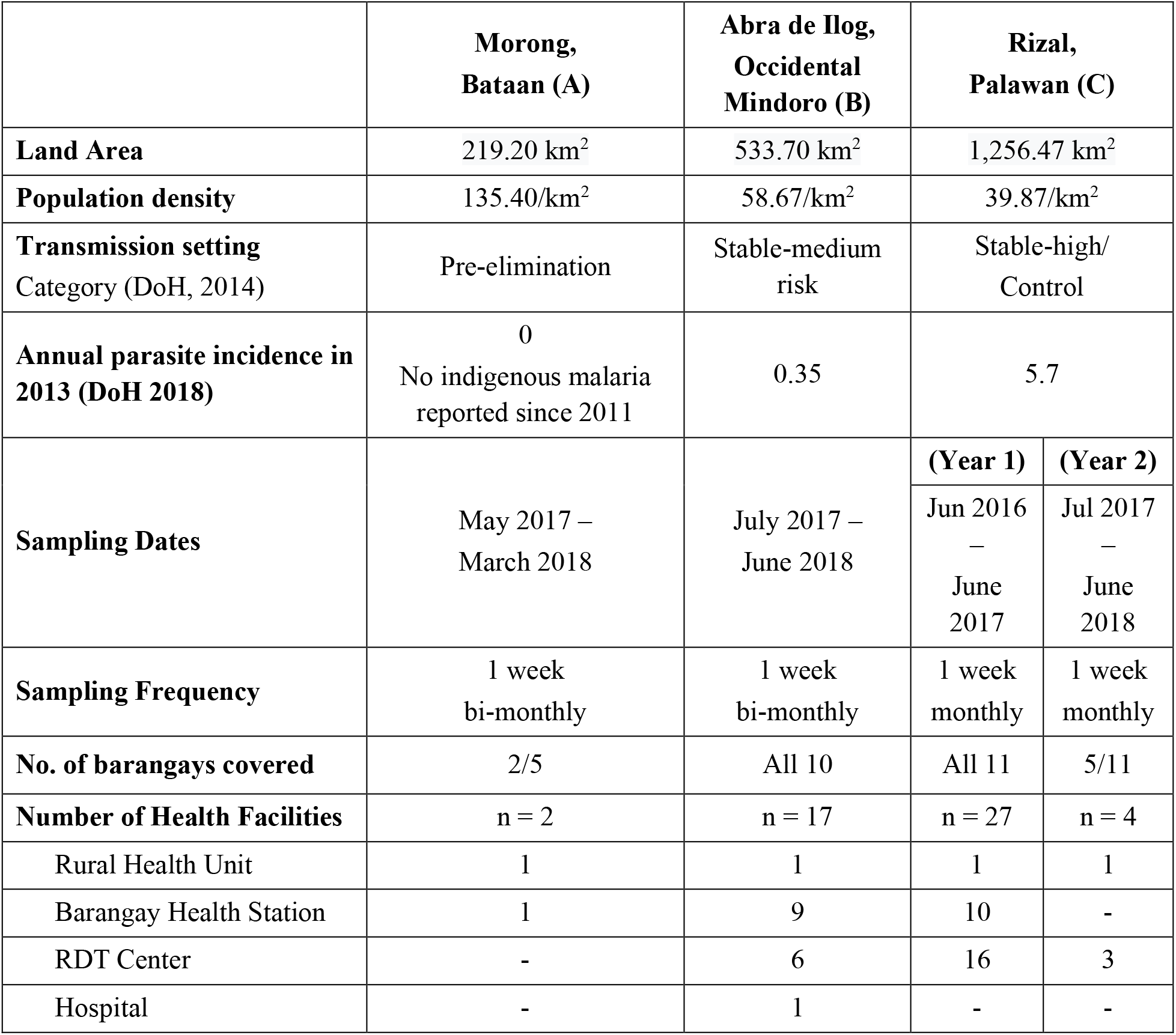
Description of study sites

**Figure 1.**
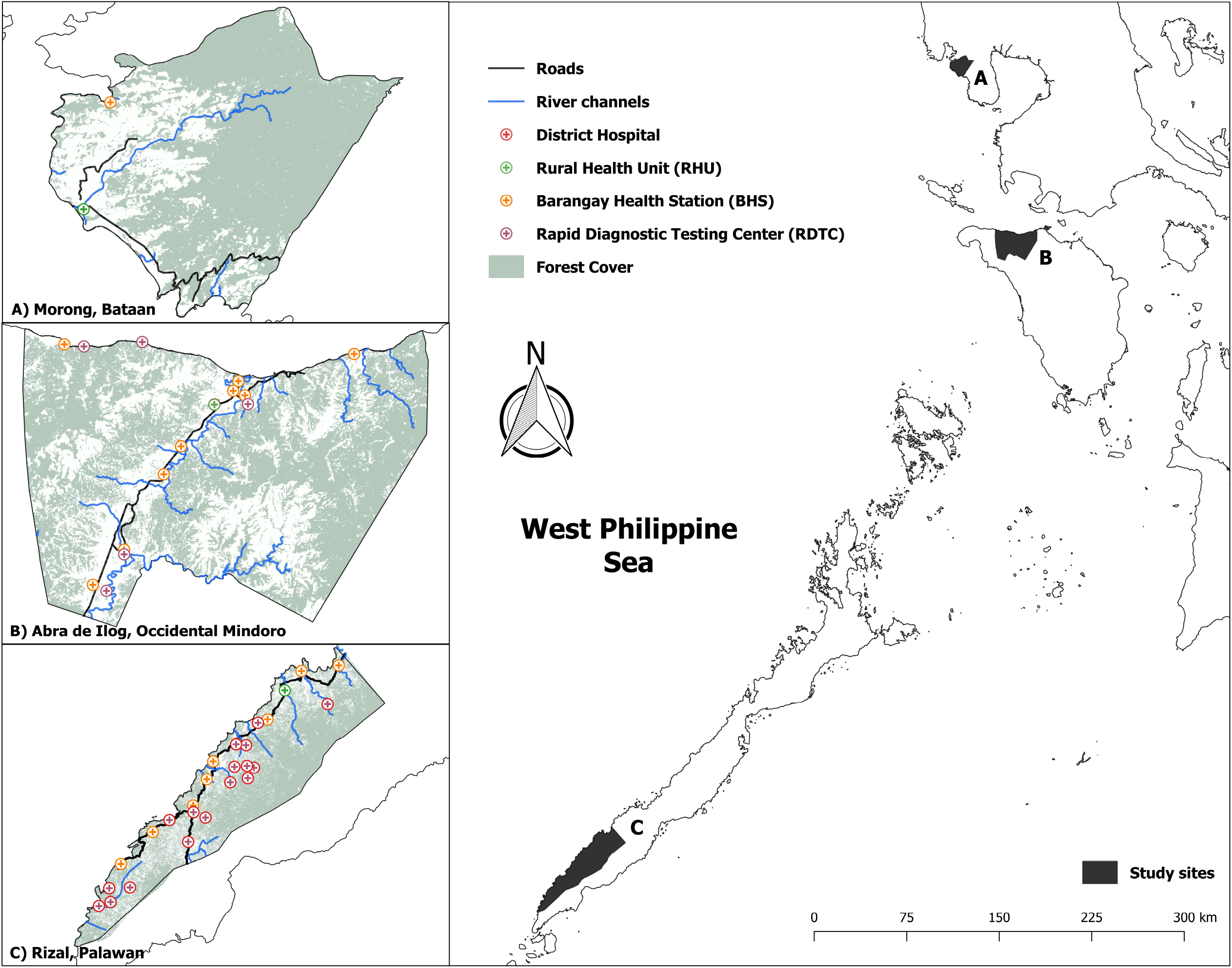
Study sites and surveyed health facilities

### Study design and sampling

Rolling cross-sectional surveys in health facilities were carried out every first week of the month for two years in Rizal. During the first year of the project (June 2016-June 2017), surveys were conducted in the 27 health facilities in the municipality (Table 1). Data collection was extended to a second year (July 2017 – June 2018), with surveys limited to the rural health center and the three malaria RDT centers that reported the highest numbers of cases the previous year. In Abra de Ilog and Morong, these surveys were conducted the first week every two months over a 12-month period. Seventeen health facilities were surveyed in Abra de Ilog. These were the rural health unit, one district hospital, nine barangay health stations and 6 RDT centers. In Morong, information was collected from the rural health unit and one barangay health station. Nearby hospitals are accessible to the residents of Abra de Ilog and Morong unlike in Rizal. Hence, residents typically opt to send their patients to these hospitals. The distance from Dr. Jose Rizal District Hospital from nearest to farthest barangay ranges from 13.7 km to 64.9 km by road. The southernmost barangays, Latud and Canipaan, were excluded as they are not accessible by road. Moreover, the Rio Tuba Nickel Mining Corporation Hospital in Bataraza, Palawan is 72.1 km away from Rizal.

Health facility staff underwent training on study procedures including obtaining written informed consent, malaria blood film and blood spot preparation, collection of geolocation information of participant’s residence, and history of illness and travel. Questionnaire data and GPS coordinates of participant households were collected using GeoODK (GeoMarvel, USA) on Android tablets using satellite imagery and known landmarks to geolocate households as described by Fornace et. al. ^10^. This included basic demographic information, symptoms, axillary temperature, movement history, malaria prevention practices and initial RDT results. Participants were classified either patient i.e. individuals seeking health consultation were referred or companions i.e. those that accompany patients. Women in the maternal clinic and individuals with serious illnesses that required urgent care or transport to higher-level health facility were excluded.

### Research Ethics

The Research Institute for Tropical Medicine – Institutional Review Board (IRB no.: 2016-04) and LSHTM (11597) approved this study.

### Assessment of malaria infection

Health facility workers collected finger prick blood samples for malaria blood film microscopy and three 20µl spots on filter paper (3MM, Whatman, Maidstone, United Kingdom). Filter papers were dried and stored with desiccant at -20 °C. Thick and thin blood films were examined by trained malaria microscopists with all positive slides and 10% of the negative slides validated by a WHO-certified level 1 malaria microscopist All participants from Rizal and Abra de Ilog were also tested for malaria using SD Bioline Malaria RDT (Abbott Rapid Diagnostics, Santa Clara, USA). All positive results from either RDT or microscopy were referred as malaria cases. Infected individuals were treated on site by the health facility personnel following the Philippines’ national treatment guidelines for malaria.

DNA was extracted from approximately 10µl of dried blood spots (DBS) on filter paper using the Chelex-100 method ^21^ modified to 6%. A nested polymerase chain reaction (PCR) assay targeting the *Plasmodium sp*. small subunit ribosomal RNA genes was used to identify genus positive species and species-specific primers were used on genus positive samples ^22, 25^. Results were visualized on a 2% agarose gel. This malaria diagnosis by PCR has a limit of detection of 0.2 parasites/uL. A subset of samples was extracted using a Qiagen DNA Mini Kit (Qiagen, Germany) to validate results. All samples were tested with PCR regardless of RDT and microscopy results; positive results were referred as malaria infections while patent infections were those individuals positive with both PCR and microscopy and/or RDT.

### Data management and analysis

Each participant was assigned a unique ID to enable linkage to samples. Data for geolocation of residence was made during the interview using designed electronic questionnaire run on GeoODK application. Participants were asked to locate their homes by pointing to its location on Android tablets. All information was later sent to the project’s secure cloud server. Households with missing GPS coordinates were visited and located using a handheld GPS (Garmin, USA) ^10^. Microscopy, RDT and PCR results were recorded in the laboratory worksheets and were double encoded using Microsoft® Excel® 2016 (Microsoft Corporation, USA) and were merged with questionnaire results. Results of malaria blood film microscopy/RDT and malaria PCR were plotted on QGIS™ Desktop software Version 3.8.2 ^26^.

All data sets were analyzed using R statistical programming language Version 3.6.3 ^27^. Individuals with incomplete outcome variables (n = 130) were excluded from analysis. For Rizal, binomial generalized mixed models were used to identify risk factors for malaria infection. An additional model was developed to determine the probability of patent infection (defined as microscopy or RDT positive infections) from all infected individuals. To select variables for inclusion, univariate analyses were conducted, with all variables with p < 0.2 screened for inclusion in multivariate analyses. The final multivariate analyses were fit in a forward-stepwise manner, with variables included in the final model with p < 0.05.

## Results

### Characteristics of study sites and population demographics

The distribution of participants by study site, nature of visit to the health facility (i.e., patient or a patient’s companion), gender, median age and presence of fever are summarized in Table 2. The majority of participants in all sites were patients rather than companions. There were higher proportions of females in all sites, with most notable difference observed in Morong, Bataan and in Abra de Ilog, Occidental Mindoro. These two sites also had much older age distributions and lower proportions of febrile individuals compared to Rizal. A review of records disclosed that in 2018, 70.8% and 61.6% of the consultations in Morong and Abra de Ilog in 2018, respectively, were for acute respiratory infections and could reflect mothers accompanying their children.

**Table 2.** Participants by province, fever and gender for all study sites

|  | Morong, Bataan | Abra de Ilog,<br>Occidental<br>Mindoro | Rizal, Palawan |  |
| --- | --- | --- | --- | --- |
|  |  |  | Year 1 | Year 2 |
| **Total Participants** | n = 896 | n = 1772 | n = 5746 | n = 1135 |
| Patients (%) | 623<br>(69.5) | 1,549<br>(87.4) | 4,391<br>(76.4) | 976<br>(86.0) |
| Companion (%) | 273<br>(30.5) | 223<br>(12.6) | 1,355<br>(23.6) | 159<br>(14.0) |
| **Fever (%)** |  |  |  |  |
| Yes (%) | 66<br>(7.4) | 76<br>(4.3) | 1,647<br>(28.7) | 406<br>(35.8) |
| No (%) | 830<br>(92.6) | 1,696<br>(95.7) | 4,071<br>(70.8) | 728<br>(64.1) |
| No data (%) | - | - | 28<br>(0.5) | 1<br>(0) |
| **Gender (%)** |  |  |  |  |
| Male (%) | 255<br>(28.5) | 617<br>(34.8) | 2,448<br>(42.1) | 528<br>(46.5) |
| Female (%) | 641<br>(71.5) | 1,155<br>(65.8) | 3,298<br>(56.8) | 605<br>(53.3) |
| No data (%) | - | - | 0<br>(1.1) | 2<br>(0.2) |
| **Age in Years, Median (IQR)** | 26<br>(11 – 39) | 28<br>(15 – 42) | 14<br>(5 – 32) | 9<br>(3 – 26) |

High proportions of health facility attendees were the Palaw’an indigenous people in both the first (63.4%, n = 3, 659) and second (46.1%, n = 523) year of surveillance in Rizal, Palawan. In contrast, clinic attendees were primarily Tagalog, the non-indigenous group, at health facilities surveyed in Abra de Ilog, Occidental Mindoro (56.4%, n = 999) and Morong, Bataan (97.3%, n = 872); while the Tagalog attendees in Palawan were 9.1% (524) in the first year and 20.2% (229) in the second year. On the other hand, only 0.4% (4) from the aboriginal group in Bataan (Aetas) and 41.6% (738) in Occidental Mindoro (Mangyans) attended the health facilities. Remaining attendees identified themselves as migrants or not originally from the province.

### Malaria Infection in patients and companions

Malaria infections were detected only in Rizal, Palawan either by RDT/microscopy or polymerase chain reaction (PCR)PCR. All samples from Abra de Ilog and Morong tested PCR-negative (Table 3). Although one RDT positive individual was detected in Occidental Mindoro, this was confirmed to be PCR-negative, suggesting a false positive RDT result or historical exposure. In the first year of collection in Rizal, there were twice the number of individuals whose PCR results were positive for malaria. It was noteworthy that 12.9% (n = 1354) of companions were positive to malaria infections by PCR contributing 28.5% (175/613) of all positive cases. PCR increased the number of participants with malaria infection in patients by 36.7% (254/693) tested by microscopy and 38% (268/706) tested by RDT. Testing by PCR and adding companions increased total infections from 6.2% (255/4095) by microscopy and 6.1% (268/4391) by RDT to 10.7% (613/5722).

**Table 3.** Malaria infection by participant category (patient or companion) for all study sites

| Study Sites | Microscopy |  | RDT |  | PCR |  |
| --- | --- | --- | --- | --- | --- | --- |
|  | + / N* | % (95% CI) | + / N* | % (95% CI) | + / N* | % (95% CI) |
| **Rizal (Year 1)** | **300 /**<br>5386 | 5.6 (5.0 –<br>6.2) | **314 /**<br>5746 | 5.5 (4.9 –<br>6.1) | **613 /**<br>5722 | 10.7 (9.9 –<br>11.5) |
| **1.23 HFs** | **176 /**<br>3922 | 4.5 (3.9 –<br>5.2) | **196 /**<br>4233 | 4.6 (4.0 –<br>5.3) | **435 /**<br>4217 | 10.3 (9.4 –<br>11.3) |
| Patient | **148 /**<br>2912 | 5.1 (4.3 –<br>5.9) | **170 /**<br>3170 | 5.4 (4.6 –<br>6.2) | **312 /**<br>3155 | 9.9 (8.9 –<br>11.0) |
| Companion | **28 / 1010** | 2.8 (1.9 –<br>4.0) | **26 /**<br>1063 | 2.4 (1.7 –<br>3.6) | **123 /**<br>1062 | 11.6 (9.8 –<br>13.6) |
| **2.4 HFs** | **124 /**<br>1464 | 8.5 (7.2 –<br>10.0) | **118 /**<br>1513 | 7.8 (6.6 –<br>9.3) | **178 /**<br>1505 | 11.8 (10.3 –<br>13.6) |
| Patient | **107 /**<br>1183 | 9 (7.5 –<br>10.8) | **98 /**<br>1221 | 8.0 (6.6 –<br>9.7) | **126 /**<br>1213 | 10.4 (8.8 –<br>12.2) |
| Companion | **17 / 281** | 6.0 (3.8 –<br>9.5) | **20 / 292** | 6.8 (4.5 –<br>10.3) | **52 / 292** | 17.8 (13.8 –<br>22.6) |
| **Rizal (Year 2; 4 HFs)** | **91 / 1102** | 8.3 (6.8 –<br>10.0) | **99 /**<br>1135 | 8.7 (7.2 –<br>10.5) | **228 /**<br>1135 | 20.1 (17.9 –<br>22.5) |
| Patient | 84 / 951 | 8.8 (7.2 –<br>10.8) | **88 / 976** | 9.0 (7.4 –<br>11.0) | **190 /**<br>976 | 19.5 (17.1 –<br>22.1) |
| Companion | **7 / 151** | 4.6 (2.3 –<br>9.3) | **11 / 159** | 6.9 (3.9 –<br>12.0) | **38 / 159** | 23.9 (17.9 –<br>31.1) |
| **Abra de Ilog** | 0 / 1640 | - | **1 / 1772** | 0.1 (0 – 0.3) | 0 / 1772 | - |
| Patient | 0 / 1427 | - | **1 / 1549** | 0.1 (0 – 0.4) | 0 / 1549 | - |
| Companion | 0 / 213 | - | 0 / 223 | - | 0 / 223 | - |
| **Morong** | 0 / 874 | - | N/A | - | 0 / 874 | - |
| Patient | 0 / 609 | - | N/A | - | 0 / 609 | - |
| Companion | 0 / 265 | - | N/A | - | 0 / 265 | - |

\*denominator for each depends on analyzable samples processed
HF's – health facilities

In the 2^nd^ year of collection, 20.1% (n = 228) of individuals were malaria positive by PCR as compared to 8.2% and 8.7% of microscopy and RDT, respectively (Table 3). Comparing the two phases of surveillance, second year of collection from the four health facilities that reported highest malaria cases confirms that proportion of PCR positives among companions (23.9%, 38/159) is high like year 1 (17.8%, 52/292) but higher compared to other facilities (11.6%, 123/1062).

Although we only sampled one week per month in Rizal, numbers of patients surveyed were 20.4% of the total patients screened by participating health facilities within an average month. Extent of coverage was highest in Taburi with 87.6% and lowest in Punta Baja with 10.5%. Coverage in other barangays ranged from 15.1% to 70.3%.

### Plasmodium species identified

Within Rizal, *P. falciparum* was the most common species detected using blood film microscopy (74.3%, 223/300) followed by *P. vivax* (18.0%, n = 54), *P. malariae* (1.3%, n = 4) and mixed infections (6.0%, n = 18); this was similar in Year 2 (76.9%, 70/91; 11.0%. n = 10 3.3%, n = 3; 4.4%, n = 4, respectively). Remaining blood films were positive for malaria but, due to poor thin smears, not speciated (Year 1, n = 1; Year 2, n = 4). By PCR, all 5 species of malaria were detected. The observations were similar with *P. falciparum* being the most prevalent species (49.9%, n = 306/613), followed by *P. vivax* (12.2%, n = 75), P. malariae (4.7%, n = 29), *P. ovale* (0.3%, n = 2), *P. knowlesi* (0.2%, n = 1) and mixed infections (8.0%, n = 49). However, 153 samples were positive of Plasmodium that were not speciated due to sample insufficiency. Likewise, PCR results in year 2 showed *P. falciparum* infection (55.3%, n = 126/228) as the most dominant species, followed by *P. vivax* (11.8%, n = 27), *P. malariae* (1.3%, n = 3) and mixed infections (9.7%, n = 22). Similar to year 1, species identification of 50 positives for *Plasmodium* were not performed (Table 4).

**Table 4.** *Plasmodium* species by malaria microscopy and PCR for Rizal

| Malaria Species | Year 1 |  |  |  | Year 2 |  |  |  |
| --- | --- | --- | --- | --- | --- | --- | --- | --- |
|  | Microscopy |  | PCR |  | Microscopy |  | PCR |  |
|  | + | % | + | % | + | % | + | % |
| *P. falciparum* | 223 | 74.3 | 306 | 49.9 | 70 | 76.9 | 126 | 55.3 |
| *P. vivax* | 54 | 18.0 | 75 | 12.2 | 10 | 11.0 | 27 | 11.8 |
| *P. malariae* | 4 | 1.3 | 29 | 4.7 | 3 | 3.3 | 3 | 1.3 |
| *P. ovale* | 0 | - | 2 | 0.3 | 0 | 0.0 | 0 | 0.0 |
| *P. knowlesi* | 0 | - | 1 | 0.2 | 0 | 0.0 | 0 | 0.0 |
| Mixed Infections | 18 | 6.0 | 49 | 8.0 | 4 | 4.4 | 22 | 9.6 |
| *Plasmodium* spp. | 1 | 0.3 | 151 | 24.6 | 4 | 4.4 | 50 | 21.9 |

### Seasonal and spatial distribution of malaria infections

For the first year of surveillance in Rizal, temporal trends of malaria infection and health facility attendance are shown in Figure 2. While there was seasonality in the numbers of patients attending health facilities and the total numbers of infections, there were some temporal trends in the proportions of individuals detected as positive by either standard or enhanced surveillance. Although the rainfall season is from May to October, increased malaria infections were only noted in the month of July and August. Similarly, second year of surveillance in Rizal focusing on health facilities with highest reported malaria cases shown some temporal trends by either surveillance method. In contrast with the first-year surveillance, malaria infections were highest in the months of February and December (Figure S1).

**Figure 2.**
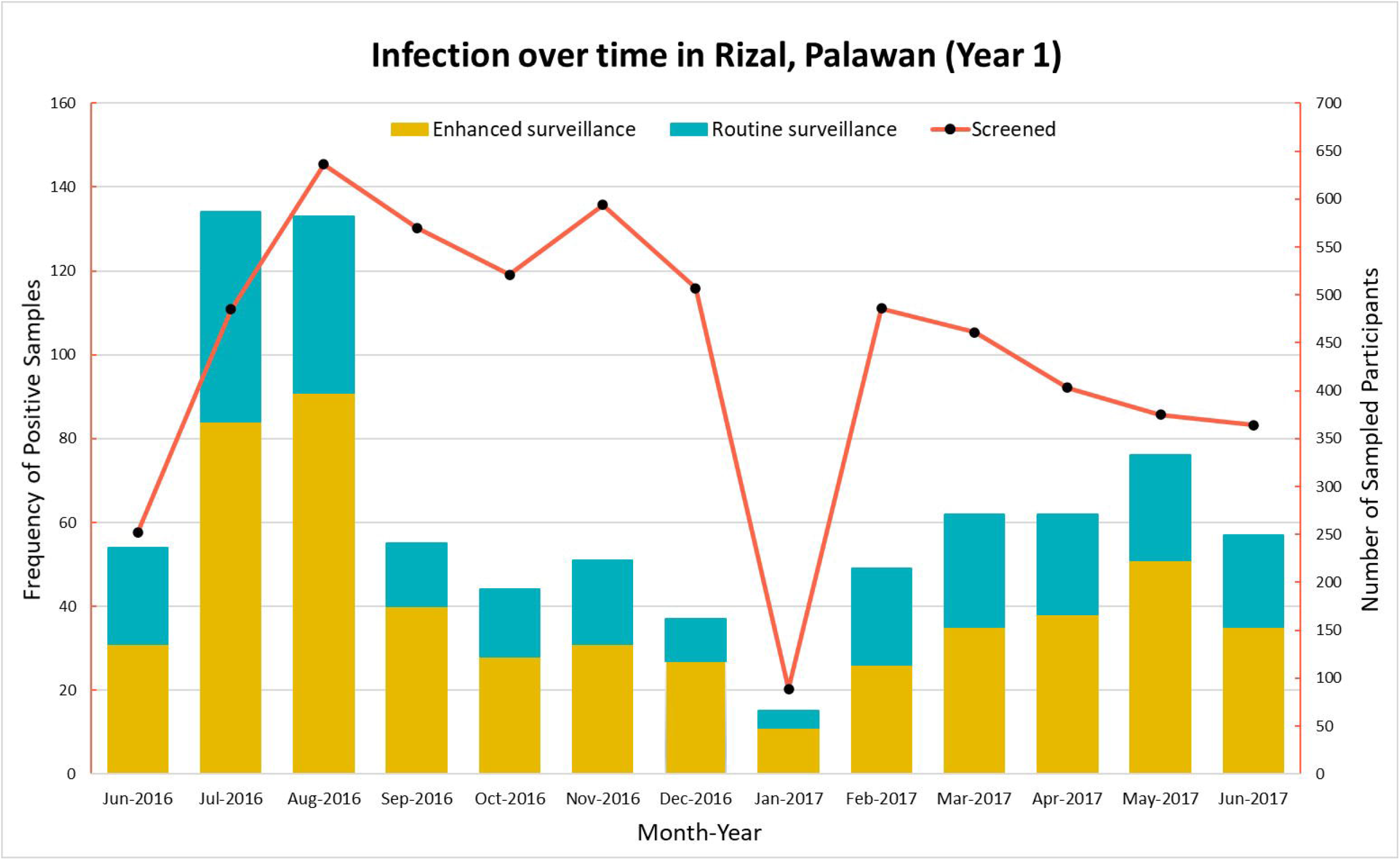
Temporal trend in Rizal, Palawan

Figure 3 shows difference in spatial distributions of infections detected by both surveillance approaches. A large proportion of infections were identified by both surveillance approaches (represented by violet points within Figure 3). While this analysis shows the utility of health facility surveys using this platform to capture real-time spatial data, analysis of spatial patterns of health facility attendance and infections were explored by Fornace, et. al, 2020 ^30^.

**Figure 3.**
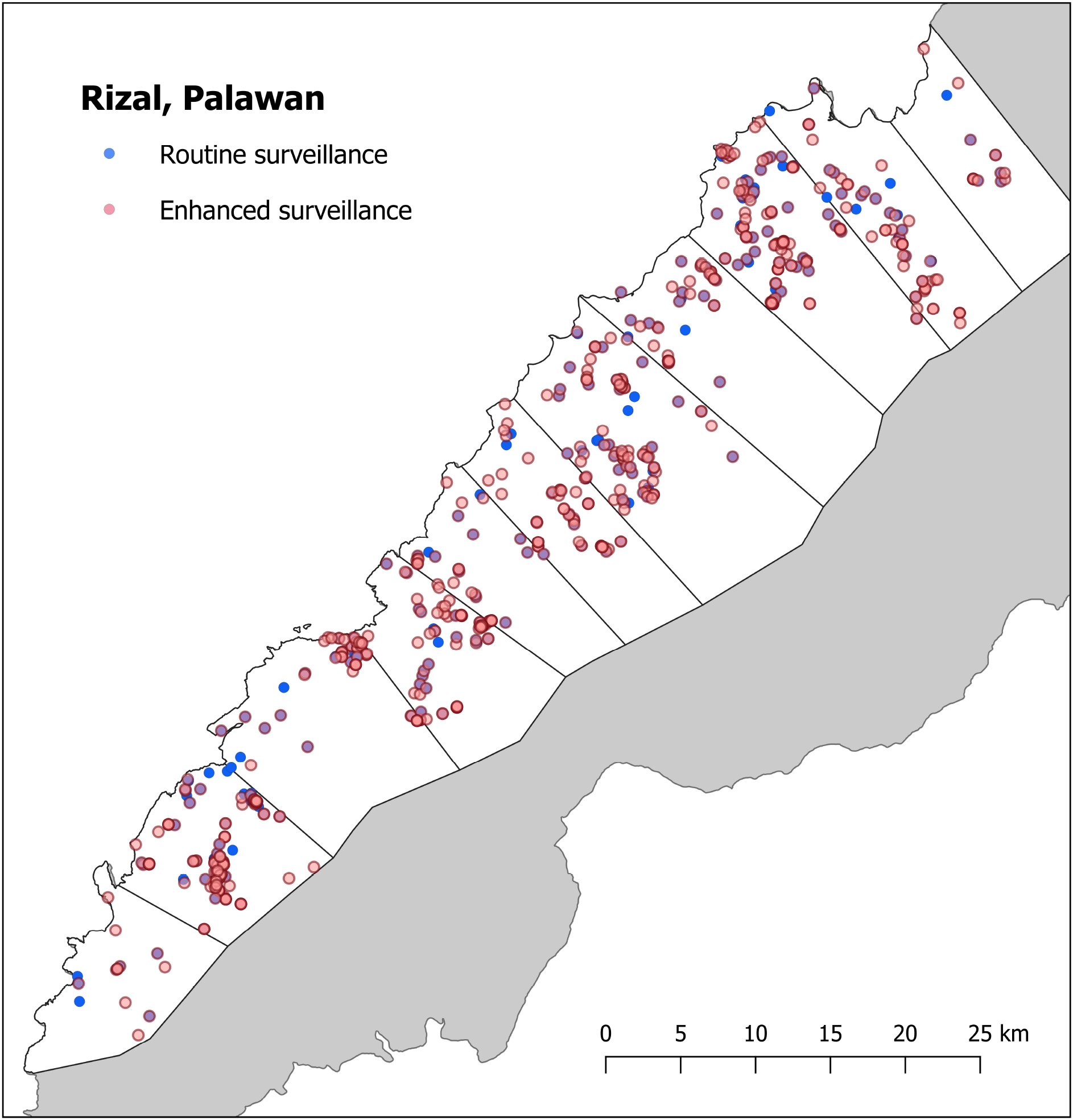
Malaria surveillance approaches

### Factors associated with malaria infections

As active malaria infections were only identified within Rizal and the first year of surveillance represented the most comprehensive dataset, we chose to focus risk factor analysis on this data. Within this year, inclusion of malaria screening of all companions increased the identification of patent infections by 16.6% (n = 60/361). This further improved to 18.5% (n = 125/676) when PCR was used to assess infection. Subsequent risk factor analysis showed that the odds of malaria infection (as detected by any diagnostic, n = 5620) were almost three times higher in 11 to 20 age group compared to over 30 years old (Table 5). Additionally, males, Palaw’an indigenous group and individuals sleeping without bednets had higher risks of infection. A significantly higher infection risk was observed in individuals with lower education levels; however, there was no clear association with specific occupational activities. There was no significant difference in infection risk detected between patients or companions screened.

**Table 5.** Risk factors for Malaria Infection in Rizal, Palawan

| Variable (n* = 5620) | UNADJUSTED |  |  | ADJUSTED |  |  |
| --- | --- | --- | --- | --- | --- | --- |
|  | OR | 95% CI | P value | OR | 95% CI | P value |
| **Age** |  |  | < 0.001 |  |  | < 0.001 |
| Under 5 | - | - |  | - | - |  |
| 5 to 10 | 1.55 | (1.19 – 2.03) |  | 2.10 | (1.56 – 2.83) |  |
| 11 to 20 | 1.59 | (1.22 – 2.07) |  | 2.64 | (1.92 – 3.64) |  |
| 21 to 30 | 1.18 | (0.87 – 1.60) |  | 1.61 | (1.15 – 2.24) |  |
| Over 30 | 0.72 | (0.54 – 0.93) |  | 0.96 | (0.72 – 1.28) |  |
| **Gender** |  |  | < 0.001 |  |  | < 0.001 |
| Female | - | - |  | - | - |  |
| Male | 1.41 | (1.19 – 1.68) |  | 1.49 | (1.24 – 1.79) |  |
| **Ethnicity** |  |  | < 0.001 |  |  | < 0.001 |
| Other Ethnicity | - | - |  | - | - |  |
| Palaw'an | 4.20 | (3.16 – 5.58) |  | 3.87 | (2.86 – 5.23) |  |
| Tagalog | 1.13 | (0.67 – 1.93) |  | 1.13 | (0.66 – 1.94) |  |
| **Occupation** |  |  |  |  |  |  |
| **1. Agriculture** |  |  | 0.079 |  |  |  |
| No | - | - |  |  |  |  |
| Yes | 0.80 | (0.63 – 1.03) |  |  |  |  |
| **2. Forestry** |  |  | 0.680 |  |  |  |
| No | - | - |  |  |  |  |
| Yes | 0.94 | (0.70 – 1.26) |  |  |  |  |
| **3. Business owner** |  |  | 0.815 |  |  |  |
| No | - | - |  |  |  |  |
| Yes | 0.94 | (0.56 – 1.58) |  |  |  |  |
| **4. Unemployed** |  |  | < 0.001 |  |  |  |
| No | - | - |  |  |  |  |
| Yes | 0.61 | (0.46 – 0.82) |  |  |  |  |
| **Activities outside house** |  |  | 0.328 |  |  |  |
| No | - | - |  |  |  |  |
| Yes | 1.09 | (0.91 – 1.31) |  |  |  |  |
| **History of travel** |  |  | 0.267 |  |  |  |
| No | - | - |  |  |  |  |
| Yes | 0.87 | (0.68 – 1.11) |  |  |  |  |
| **Type of participant** |  |  | 0.201 |  |  |  |
| Patient | - | - |  |  |  |  |
| Companion | 1.14 | (0.93 – 1.39) |  |  |  |  |
| **Education** |  |  | < 0.001 |  |  | < 0.001 |
| None | - | - |  | - | - |  |
| Primary | 0.78 | (0.65 – 0.94) |  | 0.66 | (0.53 – 0.83) |  |
| Secondary | 0.42 | (0.29 – 0.60) |  | 0.59 | (0.39 – 0.89) |  |
| **Bednet use** |  |  | < 0.001 |  |  | < 0.001 |
| Yes | - | - |  | - | - |  |
| No | 3.89 | (2.58 – 5.89) |  | 3.50 | (2.28 – 5.38) |  |
| **Health Facility Type** |  |  | 0.048 |  |  |  |
| Barangay Health Station | - | - |  |  |  |  |
| Rural Health Unit | 0.95 | (0.26 – 3.45) |  |  |  |  |
| Rapid Diagnostic Testing Center | 1.99 | (1.17 – 3.41) |  |  |  |  |

For all malaria cases, we compared the risks of patent (356/669) and subpatent malaria (313). Patent malaria infections were more common in younger age groups, with risks of patent infections decreasing with age (Table 6). Males had almost twice the odds of patent infections compared to females. Companions were more likely to have subpatent infections, as would be expected considering they were not seeking treatment. No associations between bednet use or history of travel and patent infections were identified.

**Table 6.** Patent vs Subpatent infections in Rizal, Palawan

| Variable (n* = 669) | UNADJUSTED |  |  | ADJUSTED |  |  |
| --- | --- | --- | --- | --- | --- | --- |
|  | OR | 95% CI | P value | OR | 95% CI | P value |
| **Age** |  |  | < 0.001 |  |  | < 0.001 |
| Under 5 | - | - |  | - | - |  |
| 5 to 10 | 0.81 | (0.49 – 1.18) |  | 0.86 | (0.51 – 1.46) |  |
| 11 to 20 | 0.54 | (0.19 – 0.52) |  | 0.66 | (0.40 – 1.08) |  |
| 21 to 30 | 0.28 | (0.12 – 0.39) |  | 0.43 | (0.23 – 0.80) |  |
| Over 30 | 0.24 | (0.13 – 0.61) |  | 0.29 | (0.17 – 0.50) |  |
| **Gender** |  |  | < 0.001 |  |  | < 0.001 |
| Female | - | - |  | - | - |  |
| Male | 2.24 | (1.63 – 3.09) |  | 1.99 | (1.42 – 2.79) |  |
| **Ethnicity** |  |  | 0.151 |  |  |  |
| Other Ethnicity | - | - |  |  |  |  |
| Palaw'an | 1.50 | (0.89 – 2.55) |  |  |  |  |
| Tagalog | 0.78 | (0.28 – 2.17) |  |  |  |  |
| **Occupation** |  |  |  |  |  |  |
| **1. Agriculture** |  |  | 0.015 |  |  |  |
| No | - | - |  |  |  |  |
| Yes | 0.58 | (0.37 – 0.90) |  |  |  |  |
| **2. Forestry** |  |  | 0.006 |  |  |  |
| No | - | - |  |  |  |  |
| Yes | 0.48 | (0.28 – 0.81) |  |  |  |  |
| **3. Business owner** |  |  | 0.754 |  |  |  |
| No | - | - |  |  |  |  |
| Yes | 1.16 | (0.45 – 3.02) |  |  |  |  |
| **4. Unemployed** |  |  | < 0.001 |  |  |  |
| No | - | - |  |  |  |  |
| Yes | 0.27 | (0.15 – 0.49) |  |  |  |  |
| **Activities outside house** |  |  | 0.015 |  |  |  |
| No | - | - |  |  |  |  |
| Yes | 0.67 | (0.49 – 0.93) |  |  |  |  |
| **History of travel** |  |  | 0.564 |  |  |  |
| No | - | - |  |  |  |  |
| Yes | 1.13 | (0.75 – 1.70) |  |  |  |  |
| **Type of participant** |  |  | < 0.001 |  |  | < 0.001 |
| Patient | - | - |  | - | - |  |
| Companion | 0.27 | (0.18 – 0.40) |  | 0.35 | (0.23 – 0.52) |  |
| **Education** |  |  | 0.103 |  |  |  |
| None | - | - |  |  |  |  |
| Primary | 1.05 | (0.74 – 1.48) |  |  |  |  |
| Secondary | 0.48 | (0.24 – 1.00) |  |  |  |  |
| **Bednet use** |  |  | 0.203 |  |  |  |
| Yes | - | - |  |  |  |  |
| No | 1.55 | (0.78 – 3.08) |  |  |  |  |
| **Facility Type** |  |  | 0.300 |  |  |  |
| Barangay Health Station | - | - |  |  |  |  |
| Rural Health Unit | 2.08 | (0.86 – 5.03) |  |  |  |  |
| Rapid Diagnostic Testing Center | 1.21 | (0.76 – 1.92) |  |  |  |  |

## Discussion

We developed an enhanced surveillance approach to demonstrate the utility of health facility surveys in low and high transmission settings incorporated with both molecular diagnostics and geolocation. The inclusion of companions and PCR testing provided additional information to assess transmission levels in the catchment populations that would not have been possible with the standard malaria surveillance system. The use of PCR led to an over 58% increase in the total number of infections detected from 255 by microscopy and 268 by RDT to 438. The simultaneous, collection of spatial data and use of geographic information system further increase the resolution of the spatial distribution of malaria infection. This approach can provide an operationally feasible method to supplement existing health facility data to improve surveillance and better target interventions. In areas where malaria is no longer endemic the approach provides valuable information to confirm the absence of malaria in pre-elimination settings.

By applying this approach to sites with differing transmission in the Philippines, we demonstrate how health facility surveys can complement existing malaria surveillance efforts. In the high transmission site of Rizal, we identified widespread infections in the community in addition to individuals seeking treatment. Also, with high proportion of PCR positives among companions in these health facilities compared to others, this emphasizes that these individuals must be tested especially in facilities that report high numbers of malaria. Notably, risks of infection did not differ between patients or companions, suggesting equal probabilities of infections between these two groups. This included a substantial proportion of companions who were not seeking treatment but had active febrile illnesses (26/1369). Previous studies have similarly described wider distributions of infections within populations than are captured at health facilities and highlighted the importance of identifying and targeting these infections ^13, 28, 29^. This study illustrates how screening easy access groups of health facility attendees can substantially increase the number of infections detected. By applying tablet-based applications to map the distribution of infections, this enables near real-time mapping of infections to better enable targeting of control measures ^10^.

As explored by Fornace et. al., the use of the convenience sampling of health facility attendees markedly increased detection probabilities and spatial coverage of surveillance, particularly in rural populations living in forested areas ^30^. Overall, a much wider spatial distribution of infected households was only detected by enhanced surveillance methods. Although we detected higher numbers of infections during the sampling period, this did not reflect the temporal changes of malaria throughout the year. We demonstrated the utility of this method to increase the number of infections detected but further longitudinal sampling would be required to assess fine-scale changes over time.

Additionally, we demonstrated how health facility data can be used to identify risk factors for malaria infection. Analysis of data from Rizal found risk factors for malaria infection consistent with other studies within this region, identifying higher risks in male ^31-34^ and indigenous populations ^35-38^ and individuals not using bednets ^39-41^. Although no associations were found between occupation and malaria risks, these risk factors may be partially attributed to livelihood activities such as swidden farming, movements into forested areas and associated travel and overnight stays at outdoor locations ^42-44^. As we also included molecular diagnostics in this approach, we identified significant numbers of subpatent infections, particularly in older age groups. This is consistent with other studies observing decreasing risk of patent infections with age, suggestive of acquired immunity ^45-47^. High proportions of subpatent malaria infections may contribute substantially to transmission and undermine malaria elimination efforts ^48^. This study illustrates how health facility surveys can be utilized to identify and target these infections. As this methodology collected geolocated data on use of bednets and other preventive measures as well as infection risks, this could be employed to identify priority areas for targeting control measures.

As well as identifying infections, this survey methodology allows verification of the absence of malaria transmission. Two of the study sites, Abra de Ilog and Morong, recorded no active infections. This is consistent with public health data and supports the notion that malaria transmission is all but absent in these areas. Whilst routinely collected surveillance data are key to WHO certification, augmenting these data with periodic pulses of enhanced passive or active detection provides additional assurance for the absence of infection ^49^. This can improve the statistical robustness of any assertions especially if conducted at times when historically, transmission would have been high. The use of enhanced surveys might also allow certification of elimination at lower administrative levels and assist in the more rational use of public health resources.

Despite the utility of this survey methodology, there were several important limitations to this study. This analysis relied on individuals reporting to participating health facilities and therefore is not representative of the wider population within this region. Previous studies have found biases in the demographic groups captured by facility surveys, with high attendance primarily by mothers and young children ^50^. Moreover, the indigenous populations are known to be mobile and may attend different facilities affecting the relevance of geolocation data for follow up activities. As these movements are seasonal, future studies could explore targeting specific time periods. Additionally, while the majority of infections on Rizal were *Plasmodium falciparum*, approximately a quarter were *P. vivax*; this may lead to overestimation of numbers of malaria infections if repeated reports are due to relapses. We also observed individuals (1.1 %, n = 61) who were microscopy and/or RDT positive but PCR negative. With this, there is the possibility of false-positive RDT results when the malaria parasite is cleared, and parasite antigens remain in circulation. These negative results by PCR could result from improper collection and/or storage of dried blood spots from the study sites to RITM laboratories in Manila leading to DNA degradation ^51-53^.

Nevertheless, this study demonstrates the utility of health facility surveys. Similar health facility-based approaches have been applied in Kenya ^54^, showing good concordance between facility and community-based estimates of infection. The approach has been used to identify risk factors for infection in both Haiti ^55^ and Indonesia ^10^. In this study, the addition of the combination of geolocation and diagnostic methods performed by community volunteer health workers allowed real-time mapping of field diagnostic methods such as microscopy and RDT down to household level. This is encouraging as it suggests that as strategies emerge for malaria elimination, these health workers can take new roles with proper training and resources. This is evident as they adapted to the use of mobile technology for tablet-based questionnaires and mapping and collect blood on filter paper.

## Conclusion

Extended health facility surveys can provide more comprehensive and readily accessible data for operational planning and evaluation of malaria and other diseases. Incorporating molecular diagnostics provided additional information in detecting subpatent and asymptomatic infections that are missed by routine methods such as microscopy and RDT preventing underestimated malaria prevalence. How this approach can be incorporated into routine health system and budgets requires further consideration. Community volunteer health workers can collect blood on filter paper for multiple testing or multi-disease testing in the future. Indeed, health facility surveys incorporated with geolocation and molecular methods could be adapted across range of ecologies (e.g. rural and forested population) and can support malaria control not just Palawan but other areas with similar transmission. Similarly, these methods can be used to provide stronger evidence of progress towards elimination as observed in Abra de Ilog and Morong allowing sub national verification as part of the Philippines march to malaria freedom.

## Data Availability

The datasets that support the findings of this study are available from the corresponding author on reasonable request and approval from relevant ethics committees. These are not made public due to the inclusion of identifying information on individuals that could compromise the privacy of research participants.

## Acknowledgements

The authors would like to acknowledge the Newton Fund, Philippine Council for Health Research and Development and UK Medical Research Council for funding received for ENSURE: Enhanced surveillance for control and elimination of malaria in the Philippines (MR/N019199/1). In addition, we are grateful to Ellaine Hernandez and Carol Joy Sarsadiaz for assisting field work activities of the project. Also, the local government and health staff of Rizal, Palawan for supporting the implementation of this survey.

## Disclaimer

The authors declare that they have no competing interests.

## Authors’ Addresses

Ralph A. Reyes, Research Institute for Tropical Medicine, Manila, Philippines.; Kimberly M. Fornace, London School of Hygiene and Tropical Medicine, London UK.; Maria Lourdes M Macalinao, Research Institute for Tropical Medicine, Manila, Philippines.; Beaulah L. Boncayao, Research Institute for Tropical Medicine, Manila, Philippines.; Ellaine S. De La Fuente, Research Institute for Tropical Medicine, Manila, Philippines.; Hennessey M. Sabanal, Research Institute for Tropical Medicine, Manila, Philippines.; Alison Paolo Bareng. Research Institute for Tropical Medicine, Manila, Philippines.; Inez Andrea P. Medado, Research Institute for Tropical Medicine, Manila, Philippines. Edelwisa Segubre-Mercado, Research Institute for Tropical Medicine, Manila, Philippines.; Jennifer S. Luchavez. Research Institute for Tropical Medicine, Manila, Philippines.; Julius Clemence R. Hafalla. London School of Hygiene and Tropical Medicine, London, UK.; Chris J. Drakeley. London School of Hygiene and Tropical Medicine, London UK.; Fe Esperanza Espino. Research Institute for Tropical Medicine, Manila, Philippines.

## Author contributions

FEJE, CJD, MLMM and JCRH planned and designed this study. MLMM, RAR and KMF analyzed the data. RAR, KMF, CJD and FEJE drafted the manuscript. JSL, ESM, RAR, MLMM, BLB, ESDF, HMS, APNB and IAPM supervised the data and sample collection in the study sites and analyzed samples. All authors read and approved the final manuscript.

